# Diagnostic Testing Accuracy of Sucrose-stimulated Salivary pH for Screening Early Childhood Caries Diagnosis and Severity: An Observational Study

**DOI:** 10.1101/2025.02.11.25321547

**Authors:** David Okuji, Victoria Tian, Olutayo Odusanwo, Alberta Twi-Yeboah

## Abstract

**Purpose:** The purpose of this study was to determine the diagnostic testing accuracy of child and maternal sucrose-stimulated salivary pH as screening tools for early childhood caries diagnosis and severity.

**Methods:** From a sample population of 642 mother-child dyads, with child-subjects under age six years old, child and maternal sucrose-stimulated saliva specimens were collected to measure salivary pH thirty minutes after consumption of sucrose by mouth. Immediately after the pH measurement and oral clinical examination, the pediatric dentist provider determined for child-subjects their diagnoses for early childhood caries and severity, respectively guided by the gold-standard classifications promulgated by the American Academy of Pediatric Dentistry and International Caries Detection and Assessment System. Salivary pH of 6.0 was statistically calculated as the screening cut-off point. Statistical analyses calculated values for true positive, false positive, true negative, false negative, and the diagnostic testing accuracy metrics for prevalence, specificity, sensitivity, diagnostic odds ratio, positive and negative predictive values, positive and negative likelihood ratios, and Youden’s Index. Regression models were utilized to determine the respective odds ratio associations between child and maternal sucrose-stimulated salivary pH versus the diagnosis and severity of early childhood caries.

**Results:** For early childhood caries diagnosis, child sucrose-stimulated salivary pH yielded diagnostic test accuracy metrics of 47.7 percent prevalence, 72.42 percent specificity, 59.47 percent sensitivity, 66.30 percent positive predictive value, and 66.20 percent negative predictive value, and diagnostic odds ratio 3.85. The results for maternal sucrose-stimulated salivary pH for caries severity were similar in value. Maternal sucrose-stimulated salivary pH displayed lower overall DTA metrics compared to child sucrose-stimulated salivary pH, but both child and maternal sucrose-stimulated salivary pH had higher specificity than sensitivity and displayed moderate levels of positive and negative predictive values. Child sucrose-stimulated salivary pH less than 6.0 had 3.89 times higher odds for ECC diagnosis and 3.56 times higher odds of moderate-extensive caries severity than child sucrose-stimulated salivary pH greater than 6.0. Maternal sucrose-stimulated salivary pH less than 6.0 had 1.60 times higher odds of moderate-extensive caries severity than maternal sucrose-stimulated salivary pH greater than 6.0.

**Conclusion:** Child and maternal sucrose-stimulated salivary pH screening tests are useful as a screening tool to identify children at high-risk for early childhood caries.

## INTRODUCTION

Though largely preventable, dental caries is the most common chronic disease that affects children in the U.S.^1^ The American Academy of Pediatric Dentistry (**AAPD**) defines early childhood caries (**ECC**) as the presence of one or more decayed, missing, or filled primary teeth in children aged five years or younger.^2^ More recently, the International Caries Detection and Assessment System (**ICDAS**) was developed to assess caries severity, ranging from level zero (sound tooth surface) to level six (extensive distinct cavity).^3^

ECC is characterized by its aggressive progression and ability to rapidly destroy teeth. It increases children’s risk for developing caries in their permanent dentition and for developing orthodontic problems as adults.^4^ Moreover, ECC has been shown to affect children’s quality of life beyond the oral cavity, negatively impacting nutrition, sleep, and school performance.^5^ As such, early identification of a child’s caries risk can aid in oral disease prevention and treatment planning.

Caries is a multifactorial disease: the ultimate progression of tooth decay into a cavitated lesion depends on the balance between an individual’s risk and protective factors, including diet and salivary composition. Currently, the Caries Risk Assessment (**CRA**) is a tool that takes these different factors into account to predict a patient’s likelihood of caries occurrence. While several different CRA models have been developed, there is limited scientific evidence to affirm their validity in predicting caries risk.^6,7^ Because of this, the AAPD has recommended using additional ECC risk assessment methods.^8^

Saliva has been examined as a useful alternative to assess caries risk, since it serves critical protective oral health functions of pH buffering and enamel remineralization, among others.^9^ In 1943, Dr. Robert Stephan demonstrated that after consumption of fermentable carbohydrates, acidogenic bacteria start to demineralize tooth enamel once resting salivary and plaque pH levels drop below the critical pH value of 5.5.^10^ Saliva facilitates the neutralization process that restores resting pH levels after 30 to 60 minutes and provides the source of ions that remineralize teeth after acidic encounters.^9^ Stephan reported that patients suffering from more severe tooth decay had lower resting plaque pH, as well as lower and more prolonged decreases in salivary pH after an oral glucose rinse, compared to their counterparts with less severe caries.^10^ In light of Stephan’s findings, studies have investigated salivary pH as an indicator of caries risk and found that lower salivary pH is linked to higher caries prevalence.^11–13^

In addition to child salivary pH, maternal salivary pH may be a predictor of ECC as well, due to the link between maternal and child oral health. Children born to mothers with high levels of untreated caries or tooth loss are prone to higher caries levels.^14–16^ This may be attributed in part to the vertical transmission of oral bacteria, including the highly cariogenic m*utans streptococci* (**MS**), from mother to child.^17,18^ Infants’ major source of MS is their mothers/caregivers, and mothers and infants often share the same MS genotype.^19–21^ Furthermore, accelerated maternal MS acquisition results in earlier establishment of MS in the infant oral cavity, a significant risk factor for ECC. Reduction of maternal MS levels in children younger than three years old has been shown to prevent or delay children’s MS colonization, resulting in a protective long-term effect on their risk of caries development.^22,23^

Prior research has demonstrated significant associations between both child and maternal salivary pH with ECC/Severe Early Childhood Caries (**SECC**) and caries severity, and has found that child and maternal salivary pH are positively associated with each other.^24,25^ However, studies utilizing diagnostic testing accuracy (**DTA**) methodology^26^ to explore the use of child and maternal sucrose stimulated salivary pH (**SSSpH**) as an evidence-based screening test for the diagnosis and severity of ECC are limited. Hence, utilization of implementation science principles, contextual interpretation of DTA outcomes, and provider utilization of motivational interviewing techniques can translate the DTA outcomes of an inexpensive, rapid, noninvasive, point-of-care SSSpH diagnostic screening test to serve as an early warning alert to caregivers of children.

Implementation science is defined as “the study of methods to promote the adoption and integration of evidence-based practices, interventions and policies into routine health care and public health settings.^27^” Thus, a study design which implements evidence-based science into clinical practice by using 1) a cost- and time-effective screening tool for early childhood caries diagnosis and severity, 2) contextual interpretation of the DTA outcomes from the screening tool, and 3) sharing the clinically direct DTA outcomes with the patient’s caregivers, in conjunction with motivational interviewing techniques, can demonstrate that saliva’s utility as a diagnostic screening test for childhood caries could hold significant clinical value for changing oral health behaviors, as it is an easily accessible, inexpensive, and noninvasive point-of-care diagnostic fluid.^28^

The contextual interpretation of DTA outcomes is critical to distinguish between the use of screening versus diagnostic accuracy metrics. Guidance for high value diagnostic testing suggests that the sensitivity plus specificity should be at least 1.5.^29^ Trevethan, however, asserts that for screening metrics, “sensitivity and specificity should usually be applied only in the context of describing a screening test’s attributes relative to a reference standard and predictive values are more appropriate and informative in actual screening contexts. Sensitivity and specificity can be used for screening decisions about individual people if they are extremely high. Predictive values need not always be high and might be used to advantage by adjusting the sensitivity and specificity for screening tests. In screening contexts, researchers should provide information about four metrics (sensitivity, specificity, positive predictive value, negative predictive value) and how they were derived, and clinical providers should have the skills to interpret those metrics effectively for maximum benefit to patients and the healthcare system.^30^”

Also, studies have shown that motivational interviewing techniques increase the probability that patients will improve their health behaviors, with improvements in treatment effects for the management of substance use, physical inactivity, body weight, and mortality, dental hygiene, and acceptance of further treatment and self-monitoring of health behavior for blood glucose monitoring and nutrition.^31^ In one systematic review, Rubak found that the changes in health behaviors through motivational interviewing can be indirectly measured by questionnaires (e.g., caries risk assessment (**CRA**) tools), and directly by objective measures such as blood pressure, blood glucose, weight and length of hospital stay.^32^ In a literature review Kutsch states, “The real value of pH testing may well be as a teaching tool” and “It may help (patients) in modifying behavior and choosing products that will help neutralize acid.^28^”

This study primarily aims to evaluate the DTA metrics of child and maternal SSSpH for AAPD-defined early childhood caries diagnosis and ICDAS-classified caries severity. Traditional measures of DTA, including sensitivity, specificity, positive predictive value, negative predictive value, and diagnostic odds ratio,^26^ will be calculated for both child and maternal SSSpH to assess their value as screening estimators for early childhood caries diagnosis and severity.

Secondarily, this study data will also be used to validate the associations found in previous studies between child and maternal SSSpH levels and measures of child ECC.^24,25^

## METHODS

### Study design and modeling

The study used an observational diagnostic design modeled on one of the outcomes from the classic Stephan experiment, which found that after oral exposure to simple carbohydrates, salivary and plaque pH levels decrease and gradually return to resting pH levels within thirty minutes after the exposure.^10,33–34^ Both children and their biologic mothers who met the study’s inclusion criteria were exposed to oral sucrose, and their salivary pH was measured thirty minutes after exposure. The children’s caries diagnosis and severity were classified, respectively, using the AAPD definition of early childhood caries and the ICDAS classification system for caries severity. The ability of child and maternal SSSpH to predict the child’s AAPD caries diagnosis and ICDAS caries severity was evaluated by calculating statistical DTA measures of sensitivity, specificity, positive predictive value, negative predictive value, and diagnostic odds ratio.

### Study sample size and setting

This study was approved by the NYU Grossman School of Medicine Institutional Review Board under protocol number 18-00076. A power analysis based upon the results of previous studies ^24,25^ for the association of child and maternal SSSpH for ECC, yielded a power level over 80 percent at alpha equal to 0.05, with a sample size of 119 to demonstrate a sufficient statistical power level.

An overall sample of 642 dyads of pediatric subjects and their biologic mothers were selected from the merging of two patient pool sets presenting to the dental clinic between March and October 2018 and between February 2019 and December 2020, who met the inclusion criteria at NYU Langone Hospitals-affiliated training sites in Massachusetts, Hawaii, New York, and Tennessee.

The only difference between the first and second patient pool sets, respectively, was that the first pool of subjects was instructed to refrain from brushing their teeth for 12-hours before collection of the saliva specimens and the second pool for at least 1-hour before specimen collection.

Mother-child dyads were included in the sample if the child was 0-5 years in age, categorized as American Society of Anesthesiologists (ASA) Physical Status Classification System of 1 (normal, healthy patient) or 2 (patient with mild, systemic disease),^35^ was a patient of record at an NYU Langone Hospitals-affiliated training site, had at least one erupted tooth, had a documented CRA showing caries risk, and had an oral evaluation for ECC diagnosis and ICDAS classification.

### Data collection and Quantitative variables

Resident-researchers enrolled in the NYU Langone Hospitals-Advanced Education in Pediatric Dentistry residency program performed data collection after attending a training conference to ensure one standardized protocol across different study locations. Dental caries were diagnosed via clinical and/or radiographic examinations. Following the sample selection, corresponding clinical charts were reviewed using electronic medical record software. Biologic mothers of pediatric subjects who met the inclusion criteria were invited to participate in the study and provided informed written consent to participate at the time of the visit.

Maternal- and child-subjects were instructed not to brush their teeth at least one hour prior to the sucrose challenge at the next visit, and the exact amount of time that elapsed since the last tooth brushing was recorded. The premise of this study is that variations in SSSpH may result from different stages of oral biofilm development. Subjects who last brushed their teeth less than three hours prior to saliva collection, during the initial colonization of the oral biofilm, might exhibit a different SSSpH compared to those who last brushed their teeth greater than or equal to twelve hours ago, during the rapid growth and extracellular polysaccharide production stage of the oral biofilm.^36^

At the next visit, one packet of table sugar (sucrose) was administered to the biologic mother and pediatric subject by dissolving the sugar granules on the mother’s and child’s tongue, respectively. Mother and child were asked to wait approximately 30 minutes before collecting their respective saliva specimens, and the mother was asked to complete a sociodemographic questionnaire during this 30-minute waiting period. Following the sucrose challenge, the biologic mother and child subjects were instructed to provide a stimulated saliva specimen into a cup. Study participants expectorated whole saliva into a cup for about one minute.

The resident-researcher measured the pH levels of the biologic mother’s and child’s salivary specimens by dipping pH strips, at 20-cents per strip, into the provided saliva specimens and compared immediate color change with a manufacturer-provided chart (Precision pH 4070 test strips, Precision Laboratories, Cottonwood, AZ). The child’s and mother’s pH level, quantified by the manufacturer’s strip color chart, were recorded into the child subject’s electronic dental record. Any residual saliva specimen was discarded in a sink, then flushed with water. Clinical data from the dental record, including the child’s caries diagnosis and caries severity (using the merged-ICDAS system), were abstracted after saliva specimen collection. Under the direction of the study’s Principal Investigator and local faculty mentors, resident-researchers ensured patient confidentiality, data management, and quality assurance.

The quantitative variables were defined as listed below:

- SSSpH: Measured in eight ordinal increments at pH levels 4.0, 4.4, 4.8, 5.2, 5.6, 6.0, 6.5, and 7.0.
- ECC: Defined as “the presence of one or more decayed (noncavitated or cavitated lesions), missing (due to caries), or filled tooth surfaces in any primary tooth in a child under the age of six”,^2^ as diagnosed by a dentist-provider.
- SECC: Defined as “1) any sign of smooth-surface caries in a child younger than three years of age, 2) from ages three through five, one or more cavitated, missing (due to caries), or filled smooth surfaces in primary maxillary anterior teeth, or 3) a decayed, missing, or filled score of greater than or equal to four (age three), greater than or equal to five (age four), or greater than or equal to six (age five)”^2^ as diagnosed by a dentist-provider.
- ICDAS Categories:^3^ Defined as:

- Zero = Sound tooth surface: No evidence of caries after five seconds of air drying.
- 1 = First visual change in enamel: Opacity or discoloration (white or brown) is visible at the entrance to the pit or fissure seen after prolonged air drying.
- 2 = Distinct visual change in enamel visible when wet, lesion must be visible when dry.
- 3 = Localized enamel breakdown (without clinical visual signs of dentinal involvement) seen when wet and after prolonged drying.
- 4 = Underlying dark shadow from dentine.
- 5 = Distinct cavity with visible dentine.
- 6 = Extensive (more than half the surface) distinct cavity with visible dentine.
- Time elapsed since child and biologic mother last brushed their teeth defined with the response options as:

- no response
- < 1-hour
- >= 1 < 2-hours
- >= 2 < 3-hours
- >= 3 < 4-hours
- >= 4 < 5-hours
- >= 5 < 6-hours
- >= 6 < 7-hours
- >= 7 < 8-hours
- >= 8 < 9-hours
- >= 9 < 10-hours
- >= 10 < 11-hours
- >= 11 < 12-hours
- >= 12-hours
- The diagnostic testing accuracy metrics included:^26^

- True positive (**TP**) = subjects with the disease, with the value of a parameter of interest above the cut-off
- False positive (**FP**) = subjects without the disease, with the value of a parameter of interest above the cut-off
- True negative (**TN**) = subjects without the disease, with the value of a parameter of interest below the cut-off
- False negative (**FN**) = subjects with the disease, with the value of a parameter of interest below the cut-off
- Prevalence = (TP+FN)/(TP+FN+FP+TN)
- Sensitivity (**SE**) = TP/TP+FN

- Sensitivity is defined as the probability of getting a positive test result in subjects with the disease.
- Specificity (**SP**) = TN/TN+FP

- Specificity represents the probability of a negative test result in a subject without the disease.
- Positive predictive value (**PPV**) = TP/TP+FP

- PPV represents a proportion of patients with positive test results in a total of subjects with positive result.
- Negative predictive value (**NPV**) = TN/TN+FN

- NPV is defined as a proportion of subjects without the disease with a negative test result in total of subjects with negative test results.
- Positive likelihood ratio (**LR+**) = sensitivity / (1-specificity)

- LR+ is usually higher than 1 because it is more likely that the positive test result will occur in subjects with the disease than in subjects without the disease.
- Negative likelihood ratio (**LR-**) = (1-sensitivity) / specificity

- LR-is usually less than 1 because it is less likely that negative test result occurs in subjects with than in subjects without disease.
- Diagnostic odds ratio (**DOR**) = (TP/FN)/(FP/TN)

- DOR of a test is the ratio of the odds of positivity in subjects with disease relative to the odds in subjects without disease.
- Youden’s Index (**YI**) = (sensitivity + specificity) – 1

- For a test with poor diagnostic accuracy, Youden’s index equals 0, and in a perfect test Youden’s index equals 1.

### Statistical analysis

Patient demographic and clinical characteristics were summarized in a descriptive table using mean and standard deviations for continuous variables and using counts and percentages for categorical variables. The distributions of variables were compared across groups defined by child caries diagnosis, with the two-sample independent t-test (or the ANOVA test, as appropriate) for continuous variables and the Chi-square (or the Fisher’s Exact test, as appropriate) for categorical variables. When analyzing associations with child salivary pH and fitting further regression models, the original pH levels were categorized into two levels: less than or equal to 6.0, or greater than 6.0.

The DTA metrics for prevalence, sensitivity, specificity, positive and negative predictive value, positive and negative likelihood ratio, diagnostic odds ratio, and Youden’s Index were calculated as defined above^26^ for child and maternal SSSpH versus the reference standards of prior diagnosis of ECC/SECC and prior ICDAS classification of “initial, moderate, or extensive” decay by the dental provider using oral and/or radiographic examination.

The logistic model adjusted for child age, gender, race/ethnicity, and duration of time elapsed since the last toothbrushing. Child and maternal SSSpH were used as the main prediction model.

All statistical analyses were performed in R version 4.0.3 for Mac OS.^37^ Significance level was set at 0.05.

## RESULTS

### Study population

The study included 642 pairs of child and mother subjects. Table 1 displays the overall distribution of child-level demographic and clinical characteristics, SSSpH of child and maternal saliva specimens. The pediatric subjects were 44.4 percent female, with the average age and standard deviation (SD) as 3.25 ± 1.32 years. The subjects identified ethnically as 76.5 percent non-Hispanic and racially as White (54.0 percent), Black or African American (17.8 percent), and Other, including multi-race (14.0 percent). Most subjects (96.3 percent) were defined as healthy by ASA classification, with overall prevalences showing 25.5 percent diagnosed with ECC and 21.3 percent diagnosed with SECC.

**Table 1.** Descriptive and bivariate analyses.*

| Sucrose-stimulated salivary pH (SSSpH) of Child |  |  |  |  |  |
| --- | --- | --- | --- | --- | --- |
| Variable | Variable level | Overall | SSSpH <= 6.0 | SSSpH > 6.0 | p |
| n |  | 642 | 273 | 369 |  |
| Age (years) (%) | 1-year old or less than 2-years old | 64 (10.0) | 25 ( 9.2) | 39 ( 10.6) | 0.518 |
|  | 2-years old or less than 3-years old | 131 (20.4) | 49 ( 17.9) | 82 ( 22.2) |  |
|  | 3-years old or less than 4-years old | 146 (22.7) | 64 ( 23.4) | 82 ( 22.2) |  |
|  | 4-years old or less than 5-years old | 150 (23.4) | 72 ( 26.4) | 78 ( 21.1) |  |
|  | 5-years old or less than 6-years old | 144 (22.4) | 61 ( 22.3) | 83 ( 22.5) |  |
|  | Less than 1-year old | 7 ( 1.1) | 2 ( 0.7) | 5 ( 1.4) |  |
| Age (continuous) (mean (SD)) |  | 3.25 (1.32) | 3.33 (1.28) | 3.19 (1.34) | 0.197 |
| Sex (%) | female | 285 (44.4) | 117 ( 42.9) | 168 ( 45.5) | 0.553 |
|  | male | 357 (55.6) | 156 ( 57.1) | 201 ( 54.5) |  |
| Ethnicity (%) | Hispanic | 137 (21.3) | 65 ( 23.8) | 72 ( 19.5) | 0.029 |
|  | ‡NA | 14 ( 2.2) | 10 ( 3.7) | 4 ( 1.1) |  |
|  | Non-Hispanic | 491 (76.5) | 198 ( 72.5) | 293 ( 79.4) |  |
| Race (%) | White | 347 (54.0) | 111 ( 40.7) | 236 ( 64.0) | <0.001 |
|  | Black or African American | 114 (17.8) | 62 ( 22.7) | 52 ( 14.1) |  |
|  | ‡NA | 91 (14.2) | 37 ( 13.6) | 54 ( 14.6) |  |
|  | Other, including multi | 90 (14.0) | 63 ( 23.1) | 27 ( 7.3) |  |
| **ASA Classification (%) | 1 (Normal, healthy patient) | 618 (96.3) | 260 ( 95.2) | 358 ( 97.0) | 0.173 |
|  | 2 (Patient with mild, systemic disease) | 16 ( 2.5) | 7 ( 2.6) | 9 ( 2.4) |  |
|  | ‡NA | 8 ( 1.2) | 6 ( 2.2) | 2 ( 0.5) |  |
| ***ICDAS (%) | Sound | 327 (51.3) | 91 ( 33.5) | 236 ( 64.5) | <0.001 |
|  | Initial stage decay/Moderate/severe decay | 311 (48.7) | 181 ( 66.5) | 130 ( 35.5) |  |
| Number of Child's teeth which present with the ***ICDAS Classification with greatest degree of dental caries (mean (SD)) |  | 2.29 (3.44) | 3.40 (4.08) | 1.47 (2.59) | <0.001 |
| †AAPD: Child's Caries Diagnosis (%) | Early Childhood Caries | 164 (25.5) | 93 ( 34.1) | 71 ( 19.2) | <0.001 |
|  | ‡NA | 11 ( 1.7) | 3 ( 1.1) | 8 ( 2.2) |  |
|  | No Dental Caries | 330 (51.4) | 91 ( 33.3) | 239 ( 64.8) |  |
|  | Severe Early Childhood Caries | 137 (21.3) | 86 ( 31.5) | 51 ( 13.8) |  |
| pH of child's saliva (%) | 6.5-7 | 369 (57.5) | 0 ( 0.0) | 369 (100.0) | <0.001 |
|  | 4.0-6.0 | 273 (42.5) | 273 (100.0) | 0 ( 0.0) |  |
| pH of mother's saliva (%) | 6.5-7 | 369 (57.5) | 124 ( 45.4) | 245 ( 66.4) | <0.001 |
|  | 4.0-6.0 | 273 (42.5) | 149 ( 54.6) | 124 ( 33.6) |  |
| Time between sucrose and saliva (min) (mean (SD)) |  | 32.88 (12.51) | 30.96 (13.40) | 34.30 (11.63) | 0.001 |
| Time elapsed since biologic mother last brushed her teeth (mean (SD)) |  | 7.74 (5.13) | 7.47 (5.50) | 7.95 (4.84) | 0.245 |
| Time elapsed since CHILD last had teeth brushed (mean (SD)) |  | 7.79 (5.18) | 7.50 (5.56) | 8.00 (4.88) | 0.231 |
| Sucrose-stimulated salivary pH (SSSpH) of Mother |  |  |  |  |  |
| Variable | Variable level | Overall | SSSpH <= 6.0 | SSSpH > 6.0 | p |
| n |  | 642 | 273 | 369 |  |
| Age (years) (%) | 1-year old or less than 2-years old | 64 (10.0) | 24 ( 8.8) | 40 ( 10.8) | 0.671 |
|  | 2-years old or less than 3-years old | 131 (20.4) | 60 ( 22.0) | 71 ( 19.2) |  |
|  | 3-years old or less than 4-years old | 146 (22.7) | 67 ( 24.5) | 79 ( 21.4) |  |
|  | 4-years old or less than 5-years old | 150 (23.4) | 57 ( 20.9) | 93 ( 25.2) |  |
|  | 5-years old or less than 6-years old | 144 (22.4) | 62 ( 22.7) | 82 ( 22.2) |  |
|  | Less than 1-year old | 7 ( 1.1) | 3 ( 1.1) | 4 ( 1.1) |  |
| Age (continuous) (mean (SD)) |  | 3.25 (1.32) | 3.24 (1.31) | 3.26 (1.33) | 0.848 |
| Sex (%) | female | 285 (44.4) | 115 ( 42.1) | 170 ( 46.1) | 0.36 |
|  | male | 357 (55.6) | 158 ( 57.9) | 199 ( 53.9) |  |
| Ethnicity (%) | Hispanic | 137 (21.3) | 47 ( 17.2) | 90 ( 24.4) | 0.042 |
|  | NA | 14 ( 2.2) | 4 ( 1.5) | 10 ( 2.7) |  |
|  | Non-Hispanic | 491 (76.5) | 222 ( 81.3) | 269 ( 72.9) |  |
| Race (%) | White | 347 (54.0) | 150 ( 54.9) | 197 ( 53.4) | 0.163 |
|  | Black or African American | 114 (17.8) | 41 ( 15.0) | 73 ( 19.8) |  |
|  | ‡NA | 91 (14.2) | 36 ( 13.2) | 55 ( 14.9) |  |
|  | Other, including multi | 90 (14.0) | 46 ( 16.8) | 44 ( 11.9) |  |
| **ASA Classification (%) | 1 (Normal, healthy patient) | 618 (96.3) | 262 ( 96.0) | 356 ( 96.5) | 0.013 |
|  | 2 (Patient with mild, systemic disease) | 16 ( 2.5) | 4 ( 1.5) | 12 ( 3.3) |  |
|  | ‡NA | 8 ( 1.2) | 7 ( 2.6) | 1 ( 0.3) |  |
| ***ICDAS (%) | Sound | 327 (51.3) | 128 ( 47.2) | 199 ( 54.2) | 0.096 |
|  | Initial stage decay/Moderate/severe decay | 311 (48.7) | 143 ( 52.8) | 168 ( 45.8) |  |
| Number of Child's teeth which present with the ***ICDAS Classification with greatest degree of dental caries (mean (SD)) |  | 2.29 (3.44) | 2.81 (4.00) | 1.92 (2.91) | 0.001 |
| †AAPD: Child's Caries Diagnosis (%) | Early Childhood Caries | 164 (25.5) | 67 ( 24.5) | 97 ( 26.3) | 0.393 |
|  | ‡NA | 11 ( 1.7) | 4 ( 1.5) | 7 ( 1.9) |  |
|  | No Dental Caries | 330 (51.4) | 135 ( 49.5) | 195 ( 52.8) |  |
|  | Severe Early Childhood Caries | 137 (21.3) | 67 ( 24.5) | 70 ( 19.0) |  |
| pH of child's saliva (%) | > 6.0 | 369 (57.5) | 124 ( 45.4) | 245 ( 66.4) | <0.001 |
|  | <= 6.0 | 273 (42.5) | 149 ( 54.6) | 124 ( 33.6) |  |
| pH of mother's saliva (%) | > 6.0 | 369 (57.5) | 0 ( 0.0) | 369 (100.0) | <0.001 |
|  | <= 6.0 | 273 (42.5) | 273 (100.0) | 0 ( 0.0) |  |
| Time between sucrose and saliva (min) (mean (SD)) |  | 32.88 (12.51) | 31.91 (8.91) | 33.60 (14.59) | 0.091 |
| Time elapsed since biologic mother last brushed her teeth (mean (SD)) |  | 7.74 (5.13) | 6.88 (5.05) | 8.38 (5.11) | <0.001 |
| Time elapsed since CHILD last had teeth brushed (mean (SD)) |  | 7.79 (5.18) | 6.93 (5.12) | 8.42 (5.14) | <0.001 |

\*Abbreviations used in this table. \*\*ASA = American Society of Anesthesiology.<sup>35</sup> \*\*\*ICDAS = International Caries Detection and Assessment System.<sup>3</sup> †AAPD definition of Early Childhood Caries.<sup>2</sup> ‡NA category was not considered in statistical testing.

### Descriptive and bivariate associations with child and maternal SSSpH

Child SSSpH was significantly associated with child caries diagnosis and severity (Table 1). Children with child SSSpH ≤ 6.0 were associated with a presentation of ECC (34.1 percent) or SECC (31.5 percent) at p < 0.001. They were also associated with initial, moderate, or severe tooth decay based (66.5 percent) at p < 0.001 based on ICDAS classification. A higher number of teeth with the greatest degree of caries severity by ICDAS classification was respectively associated with child SSSpH ≤ 6.0 (3.40, p < 0.001) and maternal SSSpH ≤ 6.0 (2.81, p < 0.001). As well, child SSSpH ≤ 6.0 was positively associated with maternal SSSpH ≤ 6.0 (54.6 percent, p < 0.001). Maternal SSSpH was not significantly associated with child caries diagnosis and severity. The duration of time elapsed since last brushing teeth did not contribute to any significant SSSpH differences.

### Diagnostic Testing Accuracy

Table 2 shows the DTA metrics which compare child and maternal SSSpH performance in predicting a child’s ECC diagnosis and severity relative to the reference standards of prior AAPD-defined ECC diagnosis and ICDAS-classified severity classification. Child SSSpH displayed high specificity and sensitivity in predicting both AAPD-defined ECC/SECC and ICDAS-defined caries severity. Child salivary pH had a sensitivity of 59.47 percent and specificity of 72.42 percent as a diagnostic test for caries presence. When testing for caries severity, child SSSpH demonstrated a sensitivity of 58.20 percent and specificity of 72.17 percent. The positive predictive value and negative predictive value for both dental outcomes were high and very close in value (66.30 and 66.20 percent for caries diagnosis, 66.54 and 64.48 percent for caries severity). The Diagnostic Odds Ratio (**DOR**), which compared the odds of a positive test in subjects with ECC/SECC to the odds in subjects without caries, was consistently positive (DOR = 3.85 for ECC/SECC diagnosis, DOR = 3.61 for caries severity).

**Table 2.** False Negative (FN), False Positive (FP), True Negative (TN), True Positive (TP) by child **AAPD ECC diagnosis and ***ICDAS severity gold standards.*

| Variable | Variable level | Overall (n (%)) |
| --- | --- | --- |
| n |  | 642 (100%) |
| Child- ** AAPD ECC/SECC Diagnosis | FN | 122 (19.3) |
|  | FP | 91 (14.4) |
|  | TN | 239 (37.9) |
|  | TP | 179 (28.4) |
| Child- ***ICDAS Caries Severity Diagnosis | FN | 130 (20.4) |
|  | FP | 91 (14.3) |
|  | TN | 236 (37.0) |
|  | TP | 181 (28.4) |
| Mother- ** AAPD ECC/SECC Diagnosis | FN | 147 (24.3) |
|  | FP | 135 (22.4) |
|  | TN | 188 (31.1) |
|  | TP | 134 (22.2) |
| Mother- ***ICDAS Caries Severity Diagnosis | FN | 148 (24.2) |
|  | FP | 128 (20.9) |
|  | TN | 192 (31.4) |
|  | TP | 143 (23.4) |

\*Abbreviations used in this table.
\*\*AAPD definition of Early Childhood Caries.<sup>2</sup>
\*\*\*ICDAS = International Caries Detection and Assessment System.<sup>3</sup>

**Table 3.**
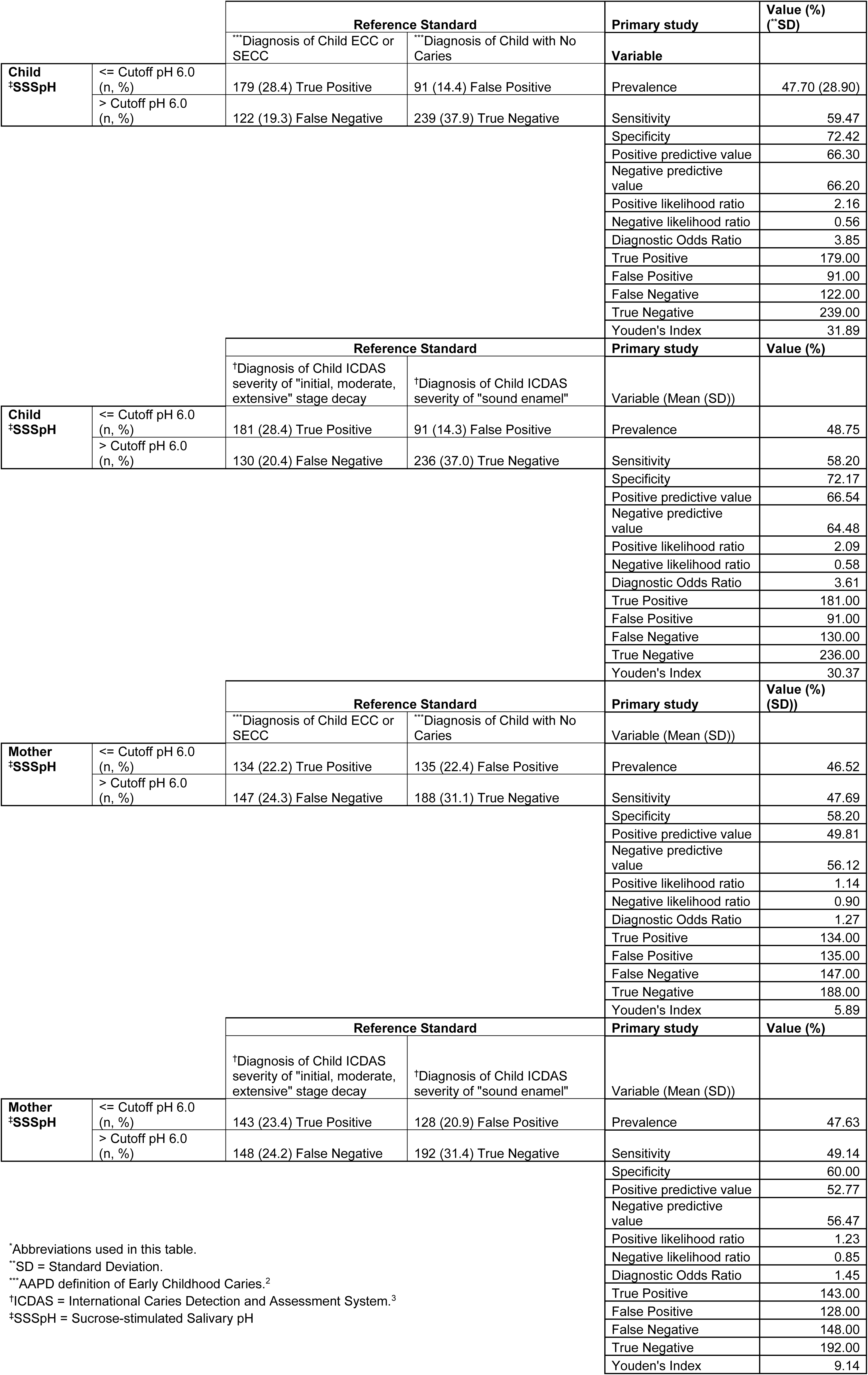
Diagnostic Test Accuracy Metrics.*

Maternal SSSpH exhibited lower sensitivity and specificity as a diagnostic test compared to child SSSpH. It had a sensitivity of 47.69 percent and specificity of 58.20 percent for AAPD-defined caries, and a sensitivity of 49.14 percent and specificity of 60.00 percent for ICDAS-classified tooth decay. Positive predictive and negative predictive values were uniformly lower as well (PPV = 49.81 percent and NPV = 56.12 percent for caries diagnosis; PPV = 52.77 percent and NPV = 56.47 percent for caries severity), and DORs were positive with lower magnitude (DOR = 1.27 for caries presence, DOR = 1.45 for caries severity).

### Logistical regression forest-plot analyses between child and maternal SSSpH and early childhood caries diagnosis and severity

The logistical regression analyses between child and maternal SSSpH and the reference standards for AAPD EEC diagnosis and ICDAS caries severity are displayed in Figure 1, respectively. Compared to those with higher SSSpH, children with SSSpH ≤ 6.0 had greater odds of having AAPD-defined ECC/SECC, ICDAS-classified tooth decay, and a higher number of teeth classified with greatest decay severity. The odds of ECC/SECC diagnosis for child subjects with SSSpH ≤ 6.0 was 3.89 [CI: 2.59, 5.90] times higher at p = 0.00 (Model 1) and the odds of having initial, moderate, or severe tooth decay was 3.56 [CI: 2.41, 5.31] times higher at p = 0.00 (Model 2) than those of children with SSSpH > 6.0.

**Figure 1.**
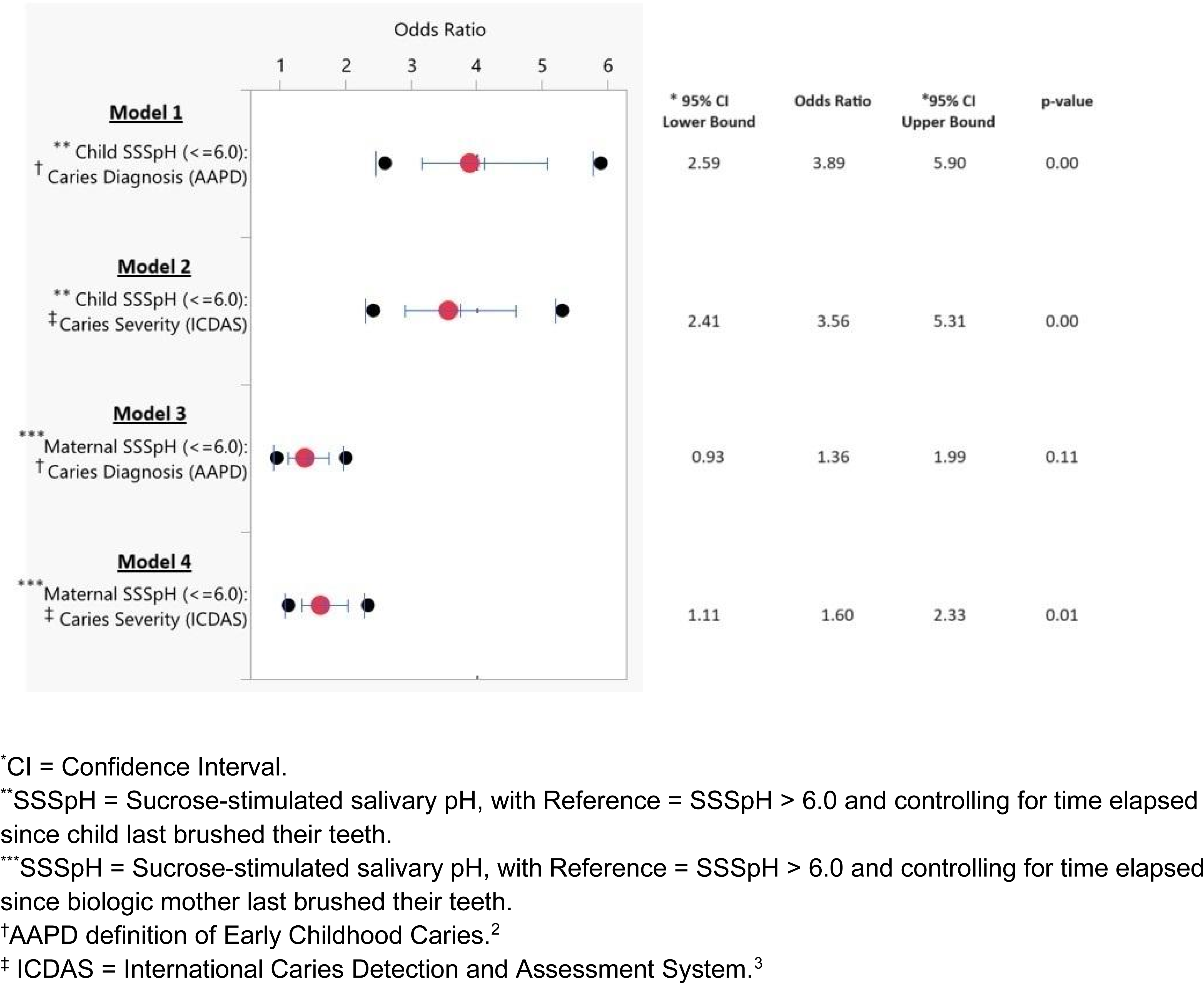
Logistical regression forest-plots for child SSSpH and maternal SSSpH less than or equal to 6.0 presenting with early childhood caries diagnosis and severity.

Models 3 and 4 present the adjusted associations between maternal subjects’ SSSpH and their children’s AAPD caries diagnosis and ICDAS caries severities, respectively. Maternal SSSpH was not significantly associated with child ECC/SECC diagnosis at p = 0.11 (Model 3). Child subjects of mothers who had SSSpH ≤ 6.0 had 1.60 times greater odds [CI: 1.11, 2.33] at p = 0.01 (Model 4) of having ICDAS-classified tooth decay compared to children of mothers with SSSpH > 6.0.

## DISCUSSION

While prior literature has investigated the association between salivary pH and child caries, and other than the lead author of this study, few if any investigators have reported DTA metrics to analyze the relationships of child and maternal SSSpH to children’s caries diagnosis and severity. Thus, this study appears to be one of the first to formally evaluate the potential of both maternal and child SSSpH as a diagnostic test for early childhood caries diagnosis and severity.

Overall, child SSSpH showed promise in identifying ECC diagnosis and severity due to its high specificity (72.42 percent for AAPD-defined caries diagnosis, 72.17 percent for ICDAS-classified caries severity) and high DOR (3.85 for ECC/SECC, 3.61 for caries severity). These metrics demonstrate that child SSSpH may be useful for categorizing children with ECC if their SSSpH ≤ 6.0. However, due to the moderate sensitivity levels (i.e., detection of false negatives), it cannot reliably assure patients they do not have ECC if their SSSpH > 6.0. Maternal SSSpH proved to be a less sensitive and less specific predictor, detecting only about half of all true positive ECC cases (47.69 percent) and slightly more than half of all true negative ECC cases (58.20 percent). In addition, lower maternal-SSSpH DOR demonstrated reduced ability to discriminate between individuals with and without ECC (DOR = 1.27 for AAPD-defined diagnosis, DOR = 1.45 for ICDAS-classified severity).

This study was also used to validate the primary author’s prior child and maternal SSSpH studies, which found that child and maternal SSSpH could signal ECC diagnosis and severity.^24,25^ It featured a larger sample size and inclusion of an additional control variable, time elapsed between last brushing and salivary testing. This study’s results provided more conclusive evidence to substantiate 1) the association between child SSSpH and ECC and 2) the association between child SSSpH and maternal SSSpH found in our prior SSSpH studies.^24,25^ However, the relationship between maternal SSSpH and ECC was weaker.

Maternal SSSpH had no significant association with ECC/SECC diagnosis and showed a reduced association with caries severity (OR = 1.60, [CI: 1.11-2.33]) Hence, the findings from this study support its primary and secondary objectives to, respectively, validate that for child and maternal SSSpH 1) DTA metrics are moderately predictive for caries diagnosis and severity and 2) support the results of previous studies which demonstrate associations between child SSSpH levels and ECC.

The results from this study are supported by previous studies. A systematic review published by Ravikumar demonstrated that children with ECC and SECC had lower salivary pH levels compared to caries-free children.^38^ Cunha-Cruz found that mean dental caries overall was 60 percent higher for individuals with a resting salivary pH ≤ 6.0 compared to those with a resting salivary pH ≥ 6.4.^39^ Choudhary found that children with decayed, missing, filled teeth (DMFT) scores of five or more showed significantly lower mean pH of resting saliva compared to children with DMFT scores less than five.^40^ Gopinath showed that children with DMFT scores greater than five had a significantly lower unstimulated salivary pH compared to those with DMFT scores equal to zero.^41^ Makawi found that the stimulated and unstimulated salivary pH in children aged 3-5 and 13-15 years with high caries risk were significantly lower compared to those with low caries risk.^42^ In a large sample size, cohort study Gao found, “a significant increase in caries risk associated with plaque acidity.^43^

Contrarily, previous studies have shown results which do not align with the present study’s findings. Jamal found no significant difference in the salivary pH in children with ECC compared to those without.^44^ In a systematic review, Khan observed that, “One study showed evidence of a protective effect of the average oral pH of stimulated salivary flow against future development of caries (Odds Ratio: 0.20).^45^”

When viewed through the lens of implementation science, contextual interpretation of DTA outcomes for screening purposes, and motivational interviewing techniques; the interpretation of the present study’s results demonstrate how screening for childhood caries and severity with child and maternal SSSpH screening tests could improve oral health behaviors and clinical outcomes.

Implementation of this inexpensive, quick, and scalable evidence-based screening test into routine health care and public health settings provides a quantitative test result to share with caregivers of children. Riley found that 73 percent of dentists only offer a qualitative caries risk assessment for children.^46^ With most dentists already performing qualitative caries risk assessments, implementation of the quantitative SSSpH screening test could easily be included in the caries risk assessment armamentarium. Also, new mothers are more likely to be receptive to ideas that would improve their offspring’s oral health, and both dental and obstetric providers have a prime opportunity to educate mothers on changes that could improve their children’s oral health.^47^ Hence, implementation of the SSSpH screening test might be well-received by caregivers of children for learning of their children’s dental caries diagnosis and severity statuses based upon quantitative and visual results.

Two independent authors, Trevethan and Power, provide guidance for contextual interpretation of DTA outcomes. Trevethan states, “A moderate PPV (with its greater proportion of false positive screening test outcomes) might be acceptable under a number of circumstances, most of which are the opposite of the situations in which a high PPV is desirable.^30^” Thus, a certain percentage of false positive outcomes might not be objectionable if follow-up tests are inexpensive, easily and quickly performed, and not stressful for patients. For example, this present study showed both child- and maternal-SSSpH displayed moderate PPV and NPV values, which limits their value in diagnostic testing but makes them preferable candidates to screen for ECC risk. Childhood caries are not contagious and generally not considered life-threatening conditions. Thus, being falsely informed of having higher caries risk (i.e., a false positive) is unlikely to harm patients or those around them and may encourage patients to engage in more protective behaviors. Furthermore, current methods for detection of early caries lesions are unreliable and often inaccurate.^48,49^ A positive SSSpH screening test may prompt dental providers to monitor a pediatric patient more proactively and motivate caregivers of children to be vigilant with dentist-prescribed oral hygiene and dietary counseling. According to Trevethan, these factors make a test with moderate PPV and NPV values acceptable for screening use.^30^

As described by Power, the “Sensitive test when Negative rules OUT the disease” (**SNOUT**) rule of thumb can be used for a screening test with high SE, low SP, moderate PPV and NPV levels, which exceed the threshold for action.^29^ The results of this study follow the independent contextual models of Trevathan and Power such that children with suspected dental caries diagnosis and severity from the child- and maternal SSSpH screening results will benefit from the dentist’s guidance for excellent oral hygiene home care combined with healthy and nutritious dietary habits.

Utilization by dentists for motivational interviewing techniques complements the contextual interpretation of the child- and maternal-SSSpH tests. Rubak suggests that clinically direct measures (e.g., SSSpH DTA), along with epidemiologically measured outcomes, ensure the reliability of the results such that the optimal study design matches the specificity and reliability of direct measures with the in-depth qualitative perspective of indirect measures (e.g., CRA tools).^32^ Hence, a simple and inexpensive SSSpH screening test for early childhood caries may help motivate parents to improve their children’s oral health outcomes.

This study’s strengths are its large sample size, inclusion of both child and maternal SSS pH measures as diagnostic predictors, and the sample population’s geographic and racial diversity, consisting of representation from four regions of the United States (Massachusetts, Hawaii, New York, Tennessee).

The study’s limitations include lack of inter-rater reliability and low socioeconomic status (**SES**) diversity. The lower SSSpH sensitivity may have been due to individual protective factors, such as salivary flow rate, buffering capacity, and oral microbiome composition, which we did not include in the study. The sample population was predominantly low SES, as many families are Medicaid eligible beneficiaries, so the results may not be generalizable to the entire population. Children of low SES families tend to have significantly worse oral health outcomes than children from moderate to high SES families.^50^

Child and maternal SSSpH tests are inexpensive, rapid, convenient, and readily scalable to identify children at high risk for ECC and promote preventive oral hygiene and nutritional behaviors.

## CONCLUSIONS

Based upon the results of this study, the following conclusions can be made: 1. Child- and maternal SSSpH screening tests are useful as a screening tool to identify children at high-risk for early childhood caries, when incorporating implementation science, contextual interpretation of diagnostic testing accuracy metrics, and motivational interviewing techniques.

## Data Availability

All data produced in the present study are available upon reasonable request to the authors.

## ACKNOWLEDGMENTS

The authors acknowledge and thank the Hansjorg Wyss Department of Plastic Surgery, NYU Langone Health for funding support; dental public health specialists Martin MacIntyre, DDS, MPH and Jay Balzer, DMD, MPH for expert review; and pediatric dental residents Mohammad Allaoa, DMD, Alexander Akbari, DDS, Tara Gainey, DDS, Anita Galloway, DDS, Johnny Joseph, DMD, Robert Kerns, DMD, MS, Spencer Kim, DDS, Kevin King, DDS, Jessica Kwon, DDS, Zohaib Munaf, DMD, MDS, Gregory Poindexter, DDS, Tashina Smiley, DMD, and Yasmina Wright, DDS for data collection contributions.

